# Efficacy, safety and dose response of STS01, a topical controlled release nanoparticle formulation (dithranol/Prosilic), in adults with mild to moderate patchy alopecia areata: A randomised, double-blind, multicentre, phase 2 trial

**DOI:** 10.64898/2026.04.02.26349934

**Authors:** Andrew G. Messenger, Alyson Bryden, Matthew J. Harries, Susan Holmes, Paul Farrant, Brian Leaker, Anita Takwale, Michelle Oakford, Manjit Kaur, Megan Mowbray, Abby E. Macbeth, Pikun Gangwani, Maria A. Gkini, Victoria M.L. Jolliffe, David Fleet

## Abstract

**Background:** There are no licensed treatments for patients with mild-to-moderate patchy alopecia areata (AA).

**Objectives:** To evaluate the efficacy, safety and dose-response of STS01, a novel nanoparticle controlled-release, topical formulation of dithranol/Prosilic.

**Methods:** In a phase 2, double-blind study, adult patients with mild-to-moderate AA (guideline ≥10% to ≤50% of scalp hair loss) were randomly assigned to STS01 at doses of 0.25%, 0.5%, 1%, 2% or placebo, daily for 6 months. The primary endpoints included the proportion of patients achieving a ≥30% improvement in Severity of Alopecia Tool (SALT) score, and percentage change from baseline in SALT score. This minimum level of improvement is generally accepted as an indicator of the population likely to progress to complete regrowth

**Results:** A total of 155 patients were randomized and treated (placebo, n=32; STS01 groups, n=30–31). STS01 1% met the primary efficacy endpoint of ≥30% SALT score improvement compared to placebo: 75.9% (95% CI, 60.3–91.4%) vs 36.7% (95% CI, 19.4–53.9%) at 6 months; p=0.0037. The least squares (LS) mean percentage change in SALT score from baseline to end-of-treatment showed a clear dose response relationship; STS01 0.5% was the minimally effective dose and 2% the maximum tolerated dose, and there was a statistically significant improvement in the STS01 1% group (−55.0% vs +0.6% with placebo; p<0.01). Significant improvements (p<0.05) in LS mean percentage changes from baseline in SALT scores were demonstrated in the STS01 1% group at 2 months (−28.6% vs 12.8%), 4 months (−57.2% vs 1.5%), and 6 months (−67.0% vs 0.6%). Clinical Global Impression-improvement was reported in 72.0% of patients with STS01 1% vs 41.7% with placebo (p<0.05). The most commonly reported treatment-emergent adverse events were skin irritation reactions, but were mostly mild (STS01: 56.7%–71.0%; placebo: 21.9%) or moderate (STS01:13.3%–35.5%; placebo: 0%) and manageable by reduced frequency of application. There were 15 skin-related discontinuations with STS01 (12.2%) and 2 (6.3%) with placebo.

**Conclusions:** STS01 demonstrated a clear dose response, with STS01 1% dose optimally more effective than placebo for hair regrowth with minimal tolerance concerns in mild-to-moderate patchy AA. Skin irritation reactions were generally manageable and there were no new safety signals. Further characterisation of the STS01 1% dose is planned in a phase 3 study.

**Lay summary:** Alopecia areata (AA) is a specific form of hair loss that occurs when the immune system attacks hair follicles causing hair to fall out. It usually starts in small, round patches. AA is a distressing condition that has many negative psychological and social effects. While there are new treatments for people with severe AA, there are still few treatments for mild/moderate patchy AA.

Dithranol is a treatment that has been used for AA. In a simple cream form it purposely irritates the skin to stimulate hair regrowth, but this also stains skin, hair and fabrics. In this study, we tested a more advanced form of dithranol, called STS01 targeting the immune system without causing irritation. STS01 has been specifically designed as a new type of cream, to reduce the irritation and staining but also improve immune response. Overall, 155 UK adults with mild/moderate patchy AA took part – one group of people were given cream that had no STS01 (placebo) and the other groups were given similar creams that contained different amounts of STS01.

We found that more participants using STS01 had hair regrowth than those using the placebo cream. Skin irritation reactions were mostly mild or moderate. Unlike previous dithranol creams, STS01 does not need to be washed off after 30 minutes, making it more convenient.

Staining of the skin occurred in very few participants. Hair regrowth was better with higher doses of STS01, but the 1% dose had fewer skin irritation reactions than the 2% dose. The 1% STS01 dose will now be tested in a larger number of people with AA to further confirm these promising results.

**What is already known about this topic?:**

- There are no licensed treatments for patients with mild to moderate patchy alopecia areata (AA), the most common form of this auto-immune condition.
- Topical dithranol is recommended on British Association of Dermatology guidelines as an irritant treatment for AA. However, these standard preparations have administration issues due to rapid oxidation, skin irritation, and staining.
- STS01 is a novel nanoparticle controlled-release once-daily topical formulation of dithranol specifically targeting down regulation of the auto-immune response and reducing administration issues

**What does this study add?:**

- This phase 2, placebo-controlled trial showed clinically relevant improvements in hair regrowth with all measures assessed.
- STS01 was generally well-tolerated with no serious adverse events; skin reactions were mostly of mild or moderate intensity.
- STS01 represents a convenient and effective approach to the treatment of mild to moderate AA.

**Graphical abstract:** 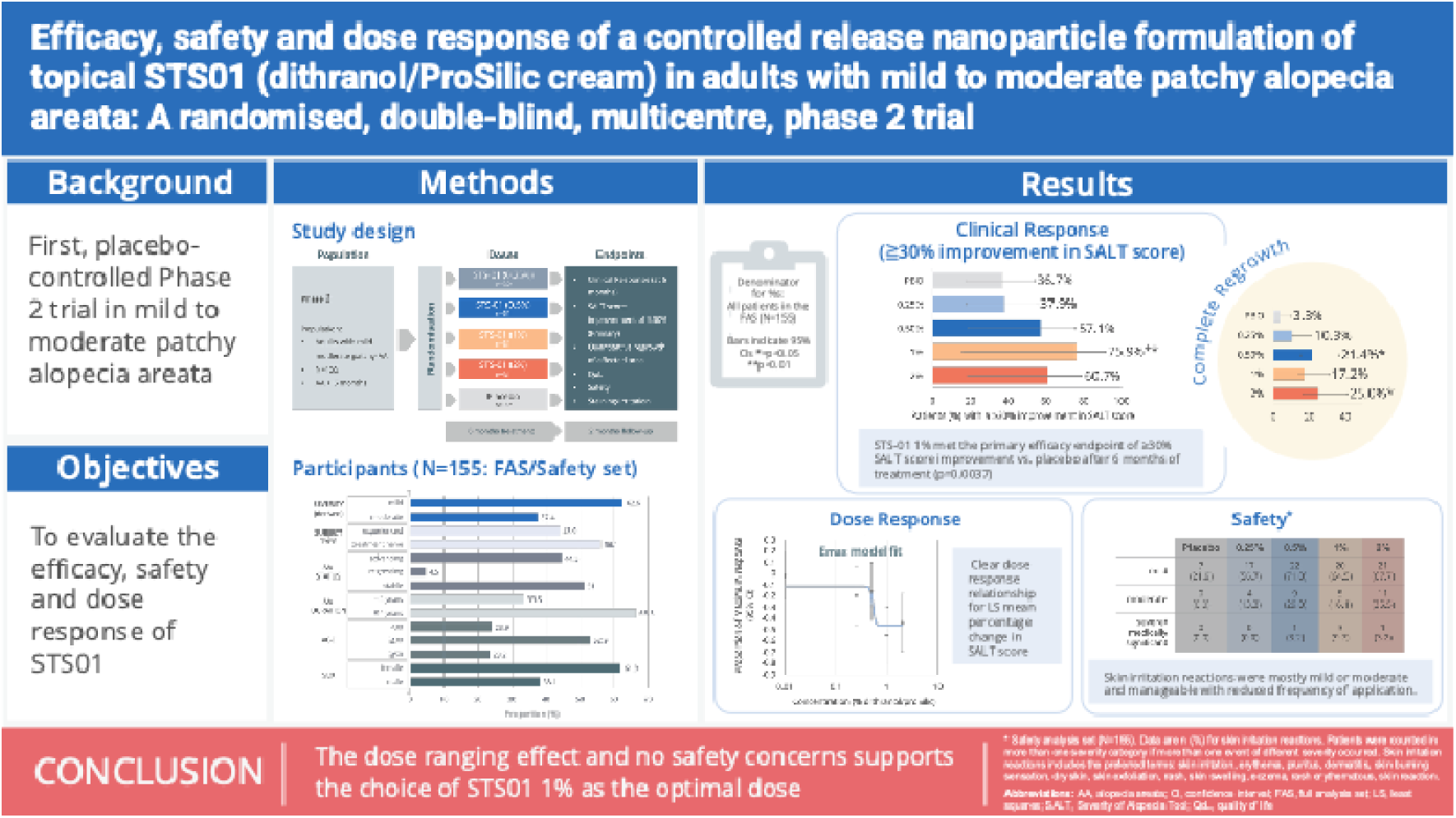

## Introduction

Alopecia areata (AA) is an auto-immune condition that leads to the sudden onset of non-scarring hair loss, with preservation of the hair follicle.^1^ AA is associated with depression and anxiety,^2,3^ a negative impact on quality of life,^4^ and an increased financial burden.^5,6^ Although over half of patients present with mild to moderate AA (i.e encompassing <50% of the scalp),^7–10^ there are no licensed treatments for these patients. JAK inhibitors are only licensed for severe AA, and they have safety warnings including for an increased risk of major cardiovascular events, venous thromboembolism and cancer.^1^ Pharmacological treatment includes topical corticosteroids, or intralesional corticosteroid injections performed by dermatologists, however evidence of their long-term efficacy is limited and there are established safety issues.^1,11^ Indeed, many people with AA experience dissatisfaction with treatment, due to lack of improvement, waning effectiveness, or side effects.^7^

Studies suggest that topical dithranol, which is indicated and was widely used for psoriasis,^12,13^ is a potentially effective treatment for AA.^14–17^ It is recommended in the British Association of Dermatology guidelines,^1^ but due to a lack of availability it is now rarely prescribed in clinical practice.^6,7^ The use of dithranol in standard formulations has been limited by several factors: it irritates skin; it stains skin, clothes, and fair hair with a purple/brown colour; application is inconvenient, particularly in the home-use setting where it is typically recommended to be washed off after 30 to 60 minutes; and it is unstable in air and sunlight, and readily oxidises to a range of products that are thought to be less active and responsible for some of these undesirable side effects.

However, dithranol has the potential to be an effective immune modifier treatment for AA if the limitations of the immediate-release formulation can be overcome. With this aim, a novel nanoparticle controlled-release, topical formulation has been developed (STS01), which combines dithranol with ProSilic®, a fully biodegradable hybrid of porous elemental silicon, lipids, and amino acids. The silicon atoms are ‘activated’ (electrically charged) to enable a controllable ‘honeycomb’ colloidal structure as a drug delivery mechanism that reduces the surface area of the drug and therefore exposure to air. Thereby enabling enhanced targeting and release characteristics.

Here we report the results of a multi-centre, double-blind, placebo controlled, phase 2 trial to assess the efficacy, safety and dose response of STS01 for the treatment of mild to moderate patchy AA.

## Methods

### Trial design and patients

The trial was conducted at 10 centres (dermatology/private clinics) in the UK from 15 March 2022 to 30 May 2024 (NCT06402630/ Eudract 2021-004145-20). The study consisted of a screening period, a 6-month double-blind treatment period, with follow up after 8 months (2 months after end of dosing).

Adults aged ≥18 years were eligible if they had a 6-month or longer history of active mild to moderate patchy AA (guideline ≥10%–≤50% of scalp hair loss), and affected skin with normal appearance. Patients were excluded if they had another type of alopecia or hair loss disorder, or any significant active scalp inflammation or condition requiring topical treatment to the scalp. Furthermore, patients were excluded if they had received any topical hair loss treatment, JAK inhibitors, systemic immunosuppressive agents, or topical calcineurin inhibitors or minoxidil medication in the previous 6 weeks, or intralesional corticosteroid medication or investigational drug treatment in the previous 3 months. These medications were also to be avoided for the duration of the study. Detailed eligibility criteria are provided in Table S1 (see Supporting Information).

### Treatment, randomization and blinding

STS01 cream is formulated in the same strengths as previously commercially available dithranol products, i.e. 0.25%, 0.5%, 1%, 2% w/w. Eligible patients were randomised to placebo or STS01 cream at the respective doses in a ratio of 1:1:1:1:1 via a centralised interactive randomisation system based on a computer-randomisation schedule with a block design that was prepared by an independent statistician.

Each dose of STS01 cream and placebo (Prosilic base control cream) were identical in colour and appearance and were supplied in identical containers and packaging. As such, all study staff and participants were blinded to treatment assignment.

Treatment was administered by the patients once daily for 6 months, at the same time each day where possible, ideally applied sparingly (completely rubbed in) in the morning. In the event of applying in the evening washing the scalp before bedtime was recommended. Treatment was continued until the end of the double-blind treatment phase, even in the event of complete hair regrowth to ensure that the condition did not return on cessation and conflict the analysis. In the event of times when administration was problematical (e.g., holidays) then omission of a few days was permitted. Following a protocol amendment, a reduction in the frequency of treatment application was permitted in patients with skin reactions. Patients were contacted one week after the start of treatment to give advice on application and on temporarily halting treatment if needed.

### Efficacy and safety endpoints and assessments

Efficacy endpoints were derived from Severity of Alopecia Tool (SALT) score ratings, which were evaluated for: clinical responder rate (i.e patients with ≥30% improvement in SALT score for the primary analysis, and ≥40%, ≥50%, ≥60%, and 100% for secondary analyses). This primary criteria was in line with other phase 2 clinical trials in severe AA where it was used as an indicator of the population most likely to completely recover hair.^18,19^ Percentage change from baseline at endpoint together with dose response relationship and percentage change over time in SALT score, with an overall risk/benefit evaluation were also performed. The SALT score ratings were performed at baseline, 2, 4 and 6 months (i.e end of treatment). All SALT scoring was performed in a blinded fashion by a single qualified central rater from random batches of images that were taken by trained site staff using a regimented protocol and standardised equipment, thereby minimising potential for between-rater variability and contribution to the margin of error.

Secondary endpoints included target-area hair growth assessment of selected AA patches with a quantitative measurement using Image J software to define hairless regions and calculate exact areas; and change in investigator-rated Clinical Global Impression (CGI)-improvement scores from baseline to the end of treatment. Treatment-emergent adverse events (TEAEs) were monitored throughout the study. A full list of endpoints is provided in the Supporting Information. These include: patient-reported outcomes (PROs) assessed using the Alopecia Areata Symptom Impact Scale (AASIS) and the Alopecia Areata Quality of Life Index (AAQLI), immunological markers (plasma cytokines) and a complete follow-up assessment 2 months post-treatment, all to be reported elsewhere.

### Statistical analysis

Taking into account that the background incidence rate of spontaneous recovery in AA symptoms is in the order of 10 to 40% depending on the severity (mild to moderate) and duration of symptoms (6 months to 2 years),^20,21^ it was determined that 147 patients would be sufficient to detect a 40% improvement in the clinical response rate or any treatment dose group vs. placebo control comparison, and a 15% dropout rate. A maximum of 25 patients per group were required for a type 1 error rate = 0.05 and 80% power.

The efficacy endpoint analyses were conducted with the full analysis set (FAS), which included all patients who applied at least one dose of medication and who subsequently provided any post baseline information. Other sensitivity analyses were performed with Per Protocol and Completers analysis sets, which confirmed and enhanced the findings with the FAS reported here. The safety analysis set included all patients who applied at least one dose of medication and considered all patients according to the treatment they actually received; with no randomisation errors, the safety set matched the FAS. All patient-reported TEAEs were assessed and reported using the safety set.

The responder rate (percentage of patients with ≥30% improvement in SALT score) was analysed using Fisher’s exact test and quantified using a binary logistic regression. Ordered categorical data were analysed using Cochran-Mantel-Haenszel (CMH – equivalent to van Elteren) tests and quantified with ordered logistic regression. Interval data were analysed using analysis of covariance (ANCOVA) models using change from baseline at 6 months (or withdrawal) as response variable, with treatment and site strata as main factors, and baseline as covariate. The SALT percentage change from baseline scores, and changes in patch area, were analysed using a repeated measures mixed model (RMMM) analysis of variance with treatment dose groups. The mixed model included treatment and site as fixed effects, baseline as a covariate, time as a repeated measure effect and patient as a random effect. The RMMM involved a reslotting strategy, and imputation for any missing visit values. All estimates of treatment effect size and the magnitude of treatment differences compared to placebo were presented as adjusted means each with 95% confidence intervals (CIs) or as odds ratios and associated p-values. Multiple comparison procedures and modelling (MCP-Mod) approach was used to assess differences between each active dose (doses categorical) and placebo and trends across the dose-response curve (doses continuous). A number of dose response models (including: linear, logistic, beta and Emax) were investigated as no prior data were available to suggest suitable fit.

The primary analysis was the SALT score ratings. Comparisons were made between each treatment dose group and placebo control. This continuous treatment effect measure (score 0 to 100) was analysed as categorical data (after responder/non-responder classification of change from baseline) and as actual continuous numeric data (actual change and percent change from baseline). The impact of dichotomising the score was investigated by examining thresholds for responder/non-responder, other specific fixed cut points of interest, and using a cumulative distribution function. Hair growth assessments were analysed using ANCOVA models. CGI-I scores were analysed using the CMH test. All evaluations were summarised descriptively.

## Results

### Patients

Overall, 185 patients were screened, 158 randomised and 155 treated, comprising the FAS and safety analysis set (Figure S1). Of the 155 treated patients, 49 (31.6%) discontinued treatment, most commonly due to loss to follow-up (n=14, 9.0%), withdrawal of consent (n=13, 8.4%), TEAEs (n=7, 4.5%), and ‘other’ (n=7, 4.5%). The results presented include all available patient data from the FAS or safety set whether they completed the trial or not.

The baseline characteristics were generally similar across treatment groups (Table 1). The median (range) age was 38.0 (18–78) years, 61.9% were women, 79.4% were White, the median duration of the current AA episode was 16 months, and 62.6% had mild scalp hair loss severity (SALT score <20).

**Table 1.** Baseline characteristics of the phase 2 population (full analysis set/safety analysis set)

|  | Placebo<br>(N=32) | STS01 cream |  |  |  | Overall<br>(N=155) |
| --- | --- | --- | --- | --- | --- | --- |
|  |  | 0.25%<br>(N=30) | 0.5%<br>(N=31) | 1%<br>(N=31) | 2%<br>(N=31) |  |
| Sex, n (%) |  |  |  |  |  |  |
| Female | 19 (59.4) | 17 (56.7) | 23 (74.2) | 16 (51.6) | 21 (67.7) | 96 (61.9) |
| Male | 13 (40.6) | 13 (43.3) | 8 (25.8) | 15 (48.4) | 10 (32.3) | 59 (38.1) |
| Age, years |  |  |  |  |  |  |
| Mean (SD) | 38.5 (12.1) | 34.2 (8.9) | 42.6 (11.5) | 37.7 (13.0) | 41.3 (12.6) | 38.9 (11.9) |
| Median (range) | 36.0<br>(18–78) | 33.5<br>(19–54) | 45.0<br>(18–64) | 37.0<br>(18–67) | 41.0<br>(19–67) | 38.0<br>(18–78) |
| Age category, years, n (%) |  |  |  |  |  |  |
| <30 | 8 (25.0) | 9 (30.0) | 5 (16.1) | 9 (29.0) | 6 (19.4) | 37 (23.9) |
| ≥30 to <50 | 18 (56.3) | 19 (63.3) | 15 (48.4) | 15 (48.4) | 15 (48.4) | 82 (52.9) |
| ≥50 | 6 (18.8) | 2 (6.7) | 11 (35.5) | 7 (22.6) | 10 (32.3) | 36 (23.2) |
| Race, n (%) |  |  |  |  |  |  |
| White | 30 (93.8) | 20 (66.7) | 26 (83.9) | 23 (74.2) | 24 (77.4) | 123 (79.4) |
| Asian-other | 1 (3.1) | 4 (13.3) | 1 (3.2) | 5 (16.1) | 3 (9.7) | 14 (9.0) |
| Black | 0 (0.0) | 3 (10.0) | 2 (6.5) | 2 (6.5) | 1 (3.2) | 8 (5.2) |
| Other | 1 (3.1) | 2 (6.7) | 1 (3.2) | 1 (3.2) | 2 (6.5) | 7 (4.5) |
| Asian-Chinese | 0 (0.0) | 1 (3.3) | 0 (0.0) | 0 (0.0) | 1 (3.2) | 2 (1.3) |
| Arab | 0 (0.0) | 0 (0.0) | 1 (3.2) | 0 (0.0) | 0 (0.0) | 1 (0.6) |
| Duration of current AA episode<sup>†</sup>, months |  |  |  |  |  |  |
|  | (n=22) | (n=22) | (n=23) | (n=22) | (n=18) | (n=107) |
| Mean (SD) | 45.9 (73.5) | 28.5 (25.9) | 28.3 (46.6) | 19.9 (16.5) | 31.8 (46.9) | 30.8 (46.2) |
| Median (range) | 22.5<br>(6–270) | 20.5<br>(6–107) | 13.0<br>(6–230) | 16.0<br>(6–72) | 10.5<br>(3–174) | 16.0<br>(3–270) |
| SALT score |  |  |  |  |  |  |
| Mean (SD) | 21.4 (20.9) | 22.3 (22.0) | 20.9 (16.9) | 19.5 (13.5) | 19.8 (15.7) | 20.7(18.1) |
| Severity<sup>‡</sup>, n (%) |  |  |  |  |  |  |
| Mild | 21 (65.6) | 19 (63.3) | 20 (64.5) | 19 (61.3) | 18 (58.1) | 97 (62.6) |
| Moderate | 11 (34.4) | 11 (36.7) | 11 (35.5) | 12 (38.7) | 13 (41.9) | 58 (37.4) |
| Type of subject, n (%) |  |  |  |  |  |  |
| Experienced | 16 (50.0) | 11 (36.7) | 14 (45.2) | 12 (38.7) | 15 (48.4) | 68 (43.9) |
| Treatment naïve | 16 (50.0) | 19 (63.3) | 17 (54.8) | 19 (61.3) | 16 (51.6) | 87 (56.1) |
| Alopecia status, n (%) |  |  |  |  |  |  |
| Advancing | 19 (59.4) | 10 (33.3) | 15 (48.4) | 17 (54.8) | 8 (25.8) | 69 (44.5) |
| Regressing | 1 (3.1) | 0 (0.0) | 0 (0.0) | 2 (6.5) | 4 (12.9) | 7 (4.5) |
| Stable | 12 (37.5) | 20 (66.7) | 16 (51.6) | 12 (38.7) | 19 (61.3) | 79 (51.0) |
| Duration of AA, n (%) |  |  |  |  |  |  |
| <4 years | 9 (28.1) | 7 (23.3) | 14 (45.2) | 12 (38.7) | 10 (32.3) | 52 (33.5) |
| ≥4 years | 23 (76.7) | 23 (71.9) | 17 (54.8) | 19 (61.3) | 21(67.7) | 103 (66.5) |

AA, alopecia areata; N, number of patients in the treatment group; n, number of patients with data; SALT, Severity of Alopecia Tool; SD, standard deviation
<sup>†</sup>In patients who provided a numerical value; patients who only provided a more general non-specific estimate of e.g < 4 years, ≥ 4 years were not included
<sup>‡</sup>Derived central SALT rater score severity grade at baseline <20=mild, ≥20=moderate

### Compliance

A precise assessment of actual use of the medication was difficult, however, the investigators reported that most patients were moderately or fully compliant (Table S2). Tubes of cream were weighed on return. The amount of cream used between visits was lower in the higher dose groups than in the placebo and lower dose groups which may be due to improvements in hair regrowth and less cream being applied (Table S2).

### Primary efficacy outcomes

After 6 months of treatment, 37.9%–75.9% of patients in the STS01 treatment groups achieved a ≥30% improvement in SALT score compared with 36.7% (95% CI, 19.4–53.9%) of patients in the placebo group, with a statistically significant difference compared to placebo in the STS01 1% group (75.9% [95% CI, 60.3–91.4%]; p= 0.0037) (Figure 1). Complete hair regrowth (100% improvement) was achieved by 10.3–25.0% of patients in the STS01 treatment groups compared with 3.3% (95% CI, 0–9.8%) in the placebo group, with statistically significant differences in the STS01 0.5% group (21.4% [95% CI, 6.2–36.6%]; p<0.05) and STS01 2% group (25.0% [95% CI, 9.0– 41.0%]; p<0.05) (Figure 1). Improvements in SALT score across a range of thresholds are shown in Table S3 and Figure 1 and demonstrate clear benefit which ever way ‘clinical improvement’ is defined.

**Figure 1.**
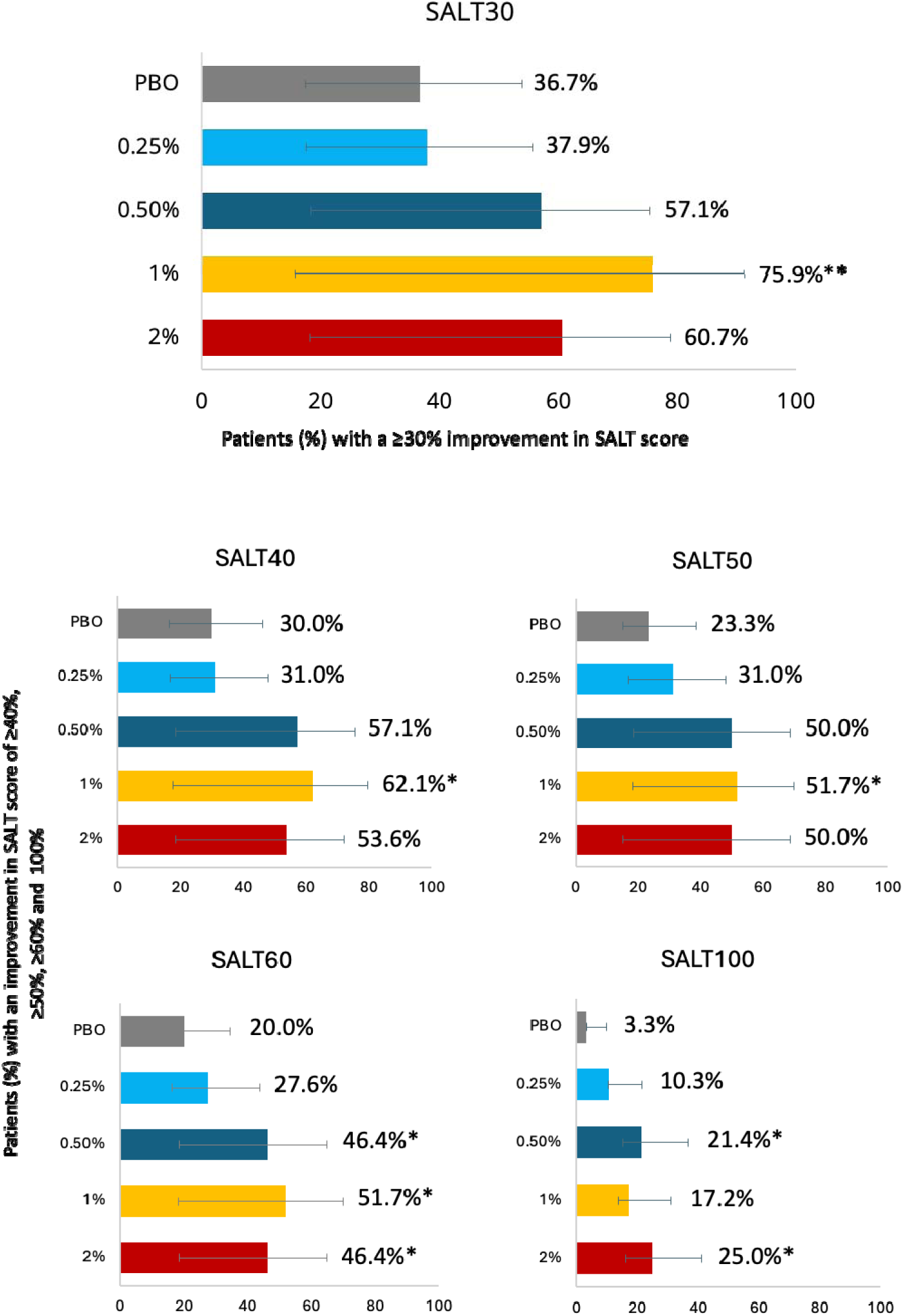
Clinical response, percentage of patients with a ≥30% (SALT30) improvement in SALT score after 6 months (primary endpoint), and a ≥40% (SALT40), ≥50% (SALT50), ≥60% (SALT60), and 100% (SALT100) improvement in SALT score after 6 months. Bars indicate 95% CIs; *=p<0.05; **p<0.01 PBO, placebo; SALT, Severity of Alopecia Tool

The LS mean percentage change from baseline in SALT score was −15.5% to −55.0% in the STS01 treatment groups, with a statistically significant −55.0% improvement in the STS01 1% group compared to a deterioration of +0.6% with placebo (p<0.01) (Table 2; Figure 2A). Similarly, the mean absolute change from baseline in SALT scores was statistically significantly improved in the STS01 1% group compared to placebo (−9.2 vs 0.7, respectively, p<0.05) (Table 2). The percentage change from baseline followed a classic dose response sigmoidal curve, with a 4 parameter Emax model proving the best fit, with an effective dose starting around 0.5% up to 2% dose strength where the dose curve plateaued (Figure 2B). STS01 1% demonstrated statistically significant improvements (p<0.05) in LS mean percentage change in SALT score from baseline at 2 months (−28.6% vs 12.8%), 4 months (−57.2% vs 1.5%), and 6 months (−67.0% vs 0.6%) (Figure 2C).

**Figure 2.**
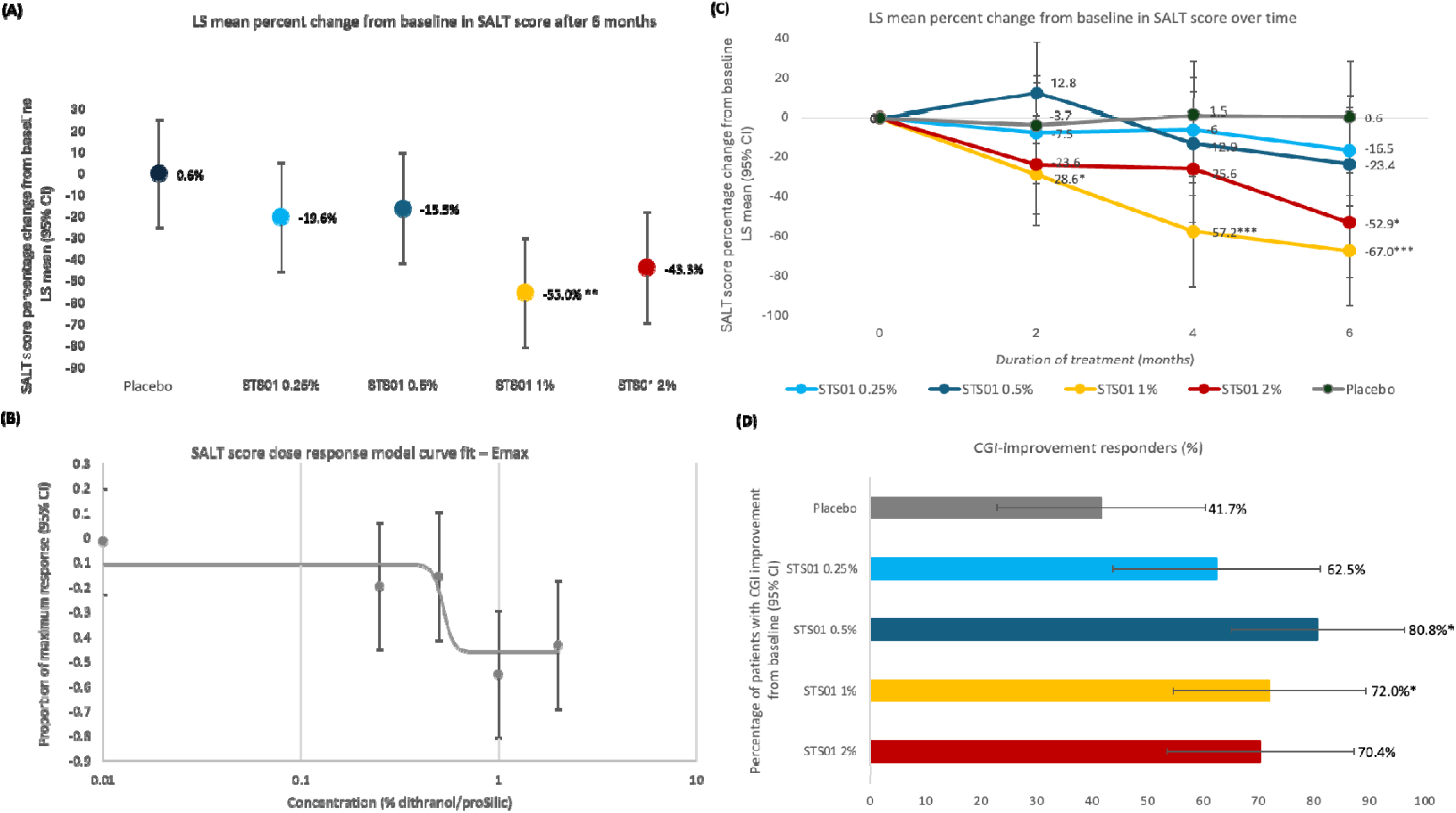
(A) SALT score percentage change from baseline (B) SALT score dose response model curve fit – Emax. (C) SALT score percentage change over time (RMMM model) (D) Investigator-assessed CGI-Improvements. Responders = percentage of patients who were assessed to be very much improved, much improved or minimally improved. Bars indicate 95% confidence intervals. CGI, Clinical Global Impression; LS, least squares; SALT, RMMM, repeated model mixed measures *=p<0.05; **p<0.01; ***p<0.001

**Table 2.** Efficacy outcomes after 6 months of treatment: Clinical responders and change from baseline in SALT score and in patch area (FAS population)

|  | Placebo<br>(N=32) | STS01 cream |  |  |  |
| --- | --- | --- | --- | --- | --- |
|  |  | 0.25%<br>(N=30) | 0.5%<br>(N=31) | 1%<br>(N=31) | 2%<br>(N=31) |
| Clinical responders: % of patients with ≥30% improvement in SALT score after 6 months |  |  |  |  |  |
| N (%) | 11 (36.7) | 11 (37.9) | 16 (57.1) | 22 (75.9)** | 17 (60.7) |
| 95% CI | (19.4, 53.9) | (20.3, 55.6) | (38.8, 75.5) | (60.3, 91.4) | (42.6, 78.8) |
| SALT score percentage change from baseline |  |  |  |  |  |
| LS mean | 0.6 | -19.6 | -15.5 | -55.0** | -43.3 |
| 95% CI | (-24.7,25.9) | (-45.2, 6.1) | (-41.5, 10.6) | (-80.7, -29.4) | (-69.3,-17.3) |
| SALT score mean absolute change from baseline |  |  |  |  |  |
| LS mean | 0.7 | 0.6 | -3.5 | -9.2* | -4.9 |
| 95% CI | (-4.3, 5.8) | (-4.5, 5.8) | (-8.7, 1.8) | (-14.4, -4.0) | (-10.1, 0.4) |
| Change from baseline in patch area for patches 1 and 2 combined (mm<sup>2</sup>)<sup>†</sup> |  |  |  |  |  |
| Patch area at baseline, mean (SD) |  |  |  |  |  |
| Patch 1 | 645.9 (340.2) | 840.3 (455.2) | 495.1 (315.1) | 697.5 (383.4) | 633.1 (400.5) |
| Patch 2 | 686.2 (512.6) | 742.7 (442.1) | 718.4 (372.3) | 666.6 (427.1) | 634.5 (400.7) |
| LS mean change from baseline (95% CI) | -116.0 (-222.7, -9.41) | -14.7 (-141.6, 112.2) | -319.6 (-450.3, -188.8) | -360.9 (-481.5, -240.2) | -163.6 (-272.0, -55.2) |
| Difference from placebo |  | 101.3 (-106.7, 309.3) | -203.5 (-415.3, 8.2) | -244.9* (-447.2, -42.5) | -47.6 (-238.8, 143.6) |
| Percent change from baseline | -6.7 (-26.9, 13.4) | -1.6 (-25.6, 22.4) | -41.5 (-66.2, -16.7) | -54.3 (-77.1, -31.5) | -12.6 (-33.1, 7.9) |
| Difference from placebo |  | 5.1 (-34.2, 44.4) | -34.7 (-74.8, 5.3) | -47.5** (-85.8, -9.3) | -5.8 (-42.0, 30.3) |

CI, confidence interval; FAS, full analysis set; LS mean, least squares mean; N, number of patients in the treatment group; n, number of patients with data at baseline and post-baseline (last observation carried forward [LOCF]); SALT, Severity of Alopecia Tool; SD, standard deviation
\*= $p < 0.05$ , \*\*= $p < 0.01$
$\geq 40$ , 50, 60, and 100% improvement in SALT score is presented in Supplementary Table S3 and Figure 1.
<sup>†</sup>If a baseline value was not recorded, the patient's screening value was used. Patients may have had only one patch. LS mean change, difference from placebo, percent change from baseline and difference from placebo refer to patches 1 and 2 combined.

### Secondary efficacy outcomes

At the last on treatment visit, AA patch areas were statistically significantly smaller with STS01 1% compared with placebo, with LS mean change from baseline of −360.9 mm^2^ (−54.3%) versus −116.0 mm^2^ (−6.7%) (p<0.05) (Table 2). The similarity in percentage changes from baseline across all the dose strengths (Table 2) reinforce the findings with primary SALT scoring undertaken from images.

According to the CGI-improvement assessment at the last on treatment visit, 62.5–80.8% of patients in the STS01 treatment groups were rated as responders (minimal, much, or very much improved) compared to 41.7% in the placebo group; the difference compared to placebo was statistically significant (p<0.05) in the STS01 0.5% and 1% groups (Figure 2D).

### Exploratory analysis

Regression analyses showed that none of the baseline demographic (age, race, sex) or clinical variables (duration of current episode of AA, severity, status, treatment status [naïve/experienced], treatment duration, site strata, duration of AA) significantly impacted the magnitude of the treatment effect on the SALT score (Table S4, Figure S2). A similar analysis with incidence and severity of irritation did not influence the magnitude of treatment outcome. This suggests the treatment effect may be quantitatively consistent across all defined patient types and does not rely on irritation to generate an efficacious response

### Safety

A total of 83.3–96.8% of patients in the STS01 treatment groups and 53.1% of patients in the placebo group experienced a TEAE (Table 3). Most the TEAEs were considered related to the treatment (66.7–96.8%). Skin irritation reactions dominated the TEAEs in the STS01 treatment groups (all skin irritation reactions: 70.0–93.5% vs 21.9% in the placebo group); the most commonly reported TEAEs (≥25% of patients) in the STS01/placebo groups, respectively, were skin irritation (38.7–54.8%/9.4%), erythema (26.7–45.2%/6.3%), pruritus (23.3–45.2%/9.4%), and dermatitis (6.7–29.0%/3.1%). Skin discolouration (staining) was reported in <=10% of patients across treatment groups. There were no serious adverse events (SAEs) and no deaths.

**Table 3.** Overview of treatment-emergent adverse events (TEAEs) (Safety set)

|  |  |  |  |  |  |
| --- | --- | --- | --- | --- | --- |
|  | Placebo | STS01 cream |  |  |  |
|  | (N=32) | 0.25%<br>(N=30) | 0.5%<br>(N=31) | 1%<br>(N=31) | 2%<br>(N=31) |
|  | Number (%) of patients |  |  |  |  |
| TEAEs | 17 (53.1) | 25 (83.3) | 29 (93.5) | 28 (90.3) | 30 (96.8) |
| Drug-related TEAEs | 8 (25.0) | 20 (66.7) | 28 (90.3) | 25 (80.6) | 30 (96.8) |
| Treatment withdrawn due to TEAE | 3 (9.4) | 4 (13.3) | 2 (6.5) | 4 (12.9) | 6 (19.4) |
| Dose reduced<sup>†</sup> due to TEAE | 0 (0.0) | 8 (26.7) | 17 (54.8) | 17 (54.8) | 21 (67.7) |
| SAE | 0 (0.0) | 0 (0.0) | 0 (0.0) | 0 (0.0) | 0 (0.0) |
| Death | 0 (0.0) | 0 (0.0) | 0 (0.0) | 0 (0.0) | 0 (0.0) |
| TEAEs by severity<sup>‡</sup> |  |  |  |  |  |
| Mild | 12 (37.5) | 23 (76.7) | 27 (87.1) | 26 (83.9) | 24 (77.4) |
| Moderate | 5 (15.6) | 8 (26.7) | 10 (32.3) | 6 (19.4) | 14 (45.2) |
| Severe | 2 (6.3) | 0 (0.0) | 1 (3.2) | 3 (9.7) | 1 (3.2) |
| Most frequently reported TEAEs (≥10 patients) |  |  |  |  |  |
| All skin irritation reactions<sup>§</sup> | 7 (21.9) | 21 (70.0) | 28 (90.3) | 24 (77.4) | 29 (93.5) |
| Skin irritation | 3 (9.4) | 15 (50.0) | 12 (38.7) | 14 (45.2) | 17 (54.8) |
| Erythema | 2 (6.3) | 8 (26.7) | 12 (38.7) | 12 (38.7) | 14 (45.2) |
| Pruritus | 3 (9.4) | 7 (23.3) | 9 (29.0) | 9 (29.0) | 14 (45.2) |
| Dermatitis | 1 (3.1) | 2 (6.7) | 9 (29.0) | 6 (19.4) | 4 (12.9) |
| Skin burning sensation | 0 (0.0) | 1 (3.3) | 3 (9.7) | 5 (16.1) | 3 (9.7) |
| Skin discolouration | 2 (6.3) | 3 (10.0) | 2 (6.5) | 3 (9.7) | 2 (6.5) |
| COVID-19 | 3 (9.4) | 1 (3.3) | 4 (12.9) | 1 (3.2) | 2 (6.5) |
| Skin irritation reactions<sup>*</sup> by severity and action taken |  |  |  |  |  |
| Severity<sup>‡</sup> |  |  |  |  |  |
| Mild | 7 (21.9) | 17 (56.7) | 22 (71.0) | 20 (64.5) | 21 (67.7) |
| Moderate | 0 (0.0) | 4 (13.3) | 9 (29.0) | 5 (16.1) | 11 (35.5) |
| Severe/medically significant | 0 (0.0) | 0 (0.0) | 1 (3.2) | 3 (9.7) | 1 (3.2) |
| Action taken |  |  |  |  |  |
| Dose reduced<sup>†</sup> | 0 (0.0) | 8 (26.7) | 17 (54.8) | 15 (48.4) | 21 (67.7) |
| Treatment withdrawn | 2 (6.3) | 4 (13.3) | 2 (6.5) | 3 (9.7) | 6 (19.4) |

SAE, serious adverse event; TEAE, treatment emergent adverse event
<sup>†</sup>Dose reduction= temporary halt of treatment<sup>‡</sup>Patients were counted in more than one severity category if more than one event of different severity occurred<sup>§</sup>Includes the preferred terms: skin irritation, erythema, pruritus, dermatitis, skin burning sensation, dry skin, skin exfoliation, rash, skin swelling, eczema, rash erythematous, skin reaction

The severity of most skin irritation TEAEs in the STS01 treatment groups was mild (56.7%–71.0%) or moderate (13.3%–35.5%), with five severe skin irritation TEAEs with STS01 (0–9.7% across groups). The proportion of patients with TEAEs leading to treatment discontinuation was 6.5–19.4% in the STS01 treatment groups vs. 9.4% in the placebo group, predominantly due to skin irritation reactions (6.5–19.4% vs. 6.3% of patients) (Table 3).

### Risk-benefit analysis

A risk-benefit analysis combining efficacy, skin reaction safety and cream application compliance endpoints demonstrated that, by 6 months, the STS01 1% dose showed clear benefits of hair regrowth outweighing skin reactions concerns even with moderate compliance (Figure S3).

## Discussion

In this phase 2, placebo controlled trial in patients with mild to moderate AA, once-daily topical treatment with STS01, a controlled release, topical formulation of dithranol, met the primary efficacy endpoint of ≥30% SALT score improvement compared to patients receiving placebo. The improvements were consistent across all the clinical measures assessed including other clinical improvement criteria (≥40, ≥50, ≥60 and 100%) which were in-line with the criteria used at phase 2 in severe AA with barictinib and ritlecitinib.^18,19^ The response profiles over time also showed that observed improvements at the 2-month visit continued to increase over the 6-month treatment period and furthermore, during this period STS01 was generally well-tolerated with respect to reported skin irritation reactions and no unexpected adverse events. This is the first formal phase 2 randomised double blind and placebo-controlled investigation to assess a treatment in patients with mild or moderate patchy AA, currently an area of substantial unmet need with no licenced treatments. These results show that STS01 represents a simple, convenient and effective approach to the treatment of the mild to moderate AA condition.

Although our results demonstrate clear evidence of clinical benefit which reinforces a possible down regulation of the immune system as the major mechanism for regrowth in AA. It is suggested that this involved modification of cytokines. promoted by epidermal T cell lymphocytes and disruption of cell function leading to anti-proliferation of the inflammatory cell infiltrate in the AA-impaired hair follicle.^22–26^. This subject is addressed, specifically with dithranol, in a subsequent publication.

It is well documented that increasing doses of dithranol (0.1 to 3.0%) in conventional formulations have a close relationship with the extent of skin erythema and staining.^27^ These adverse effects result from immediate release of dithranol and an oxidative reaction in air when applied, resulting in large amounts of the dimer degradative product, which causes visible brown staining. In our investigation, very little difference was observed between any of the four STS01 doses with respect to incidence and severity of skin irritation reaction TEAEs and with few reports of skin staining (10/123 patients who received STS01). This finding supports the successful achievement of the control of drug release and permeation into the surface of the affected scalp with the ProSilic formulation and suitability for once daily application. Controlling the release of dithranol has been shown previously to have a positive impact on irritation and staining in other skin model experiments.^28^

As dithranol is a known irritant, some erythema was not unexpected, especially in sensitive skinned patients, using this new formulation. Most irritation was of mild to moderate intensity (across all doses) and in the limited instances where a severe reaction occurred clinical advice to temporarily halt treatment (termed dose reduction) aided management of the event. Only 9.7% patients in the 1% STS01 group discontinued due to these reactions and although 19% of the STS01 2% dose group had the dose withdrawn, 25% actually achieved complete regrowth confirming a possible improved efficacy if tolerance is acceptable.

The STS01 controlled release formulation addresses the opportunity to apply the cream in the morning without the need to wash off later as with standard dithranol preparations. Therefore, the formulation offers a more tolerable option with regards to skin irritation, as well as being more convenient with regard to home application and staining.

Dithranol has been commonly used for the treatment of psoriasis for several decades, with no major safety concerns,^12,13^ and with no new safety signals emerging in the trial, STS01 appears to have an acceptable safety profile. A phase 3 trial, evaluating the efficacy and safety of the 1% formulation of STS01 is now planned. The 1% dose has been chosen based on the risk-benefit analysis showing the best balance between efficacy, safety and compliance.

In conclusion, the novel formulation of dithranol demonstrated a clear dose response in this phase 2 trial, with STS01 1% optimally more effective than placebo for hair regrowth and minimal tolerance concerns in mild to moderate patchy AA. Skin irritation reactions were generally manageable and there were no new safety signals. Further characterisation of the efficacy and safety of the 1% dose is now planned in a phase 3 clinical study.

Following on from this report further investigations of: Immune mechanisms permitting hair regrowth, SALT scoring methodology and Quality of Life implications with STS01 treatment will also be published.

## Acknowledgements

We are grateful to the patients and the clinical trial staff for their participation in the study. Under the direction of the authors, Amanda Prowse PhD of Lochside Medical Communications (Glasgow, UK) provided medical writing, which was funded by Soterios Ltd, in accordance with Good Publication Practice guidelines. Support for the graphical abstract was provided by Emma Melchor PhD (Emma Melchor Illustration, Glasgow, UK), funded by Soterios Ltd. MJH is supported by the NIHR Manchester Biomedical Research Centre (NIHR203308).

## Funding source

This research was funded by Soterios Ltd, UK. The trial sponsor was involved in the study design, the collection, analysis, interpretation of data and the decision to submit the article for publication. Soterios Ltd also paid for professional writing assistance and article processing charges.

## Conflicts of interest

Chief Investigator **AGM** reports fees from Soterios Ltd. Chief Statistician **DMF** is an employee of Soterios Ltd.

All other authors were Principal Investigators in the trial and their clinics were reimbursed for the work involved. Most also had sponsorship in the form of consultancies, investigational roles or lecturing roles on behalf of other Dermatological pharmaceutical companies

## Data availability

The data underlying this article will be shared upon reasonable request to the corresponding author.

## Ethics statement

The trial was conducted in accordance with the ethical principles that have their origins in the Declaration of Helsinki and International Council for Harmonisation (ICH) Good Clinical Practice (GCP). The responsible Independent Ethics Committee (London - South East Research Ethics Committee) approved the study protocol (Reference: 21/LO/0851).

## Patient consent

All patients provided written informed consent.

## Supplementary information

### Patient disposition

**Figure S1.**
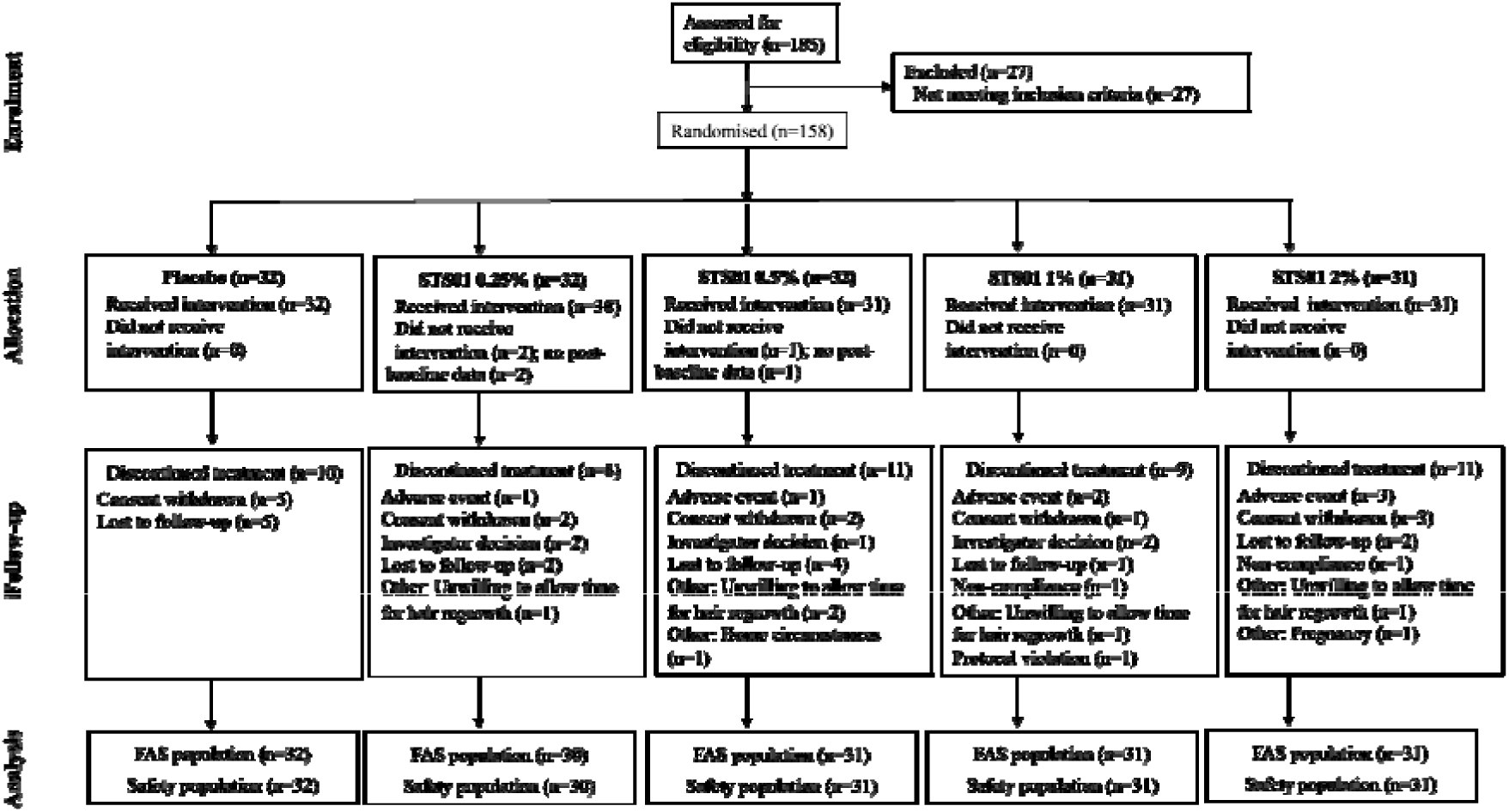
Patient flow diagram (CONSORT)

### Eligibility

**Table S1.** Eligibility criteria.

| Inclusion criteria |
| --- |
| Men or women $\geq 18$ years, with a diagnosis of mild to moderate patchy AA. |
| No more than 50% loss of scalp hair (≤SALT 50), and no less than 10% (≥SALT 10), although investigator discretion was permitted if at least one distinct patch was visible. Evidence of active disease, in the judgement of the physician, was also required. Active disease was indicated by the presence of bald patches devoid of hair and with no evidence of hair regrowth. The presence of exclamation mark hairs and a positive pull test at the margins were further indicators of disease activity but were not essential |
| Active AA present for at least 6 months |
| Affected skin with normal appearance and no grossly evident epidermal alterations such as scaling or follicular abnormalities. |
| General health status acceptable for participation in a 6-month clinical trial. |
| Women of childbearing potential had a negative urine pregnancy test |
| Exclusion criteria |
| Any diagnosed other types of alopecia (including alopecia areata totalis, alopecia areata universalis and diffuse alopecia areata). Ophiasis was acceptable if the original hair line was evident and, in the opinion of the investigator, a SALT score could be calculated reliably. Where ophiasis was extensive, e.g., involving the entire occipital hairline, and/or the position of the pre-morbid hairline was uncertain, the patient was excluded. |
| Other hair loss disorders e.g., scarring alopecia (except androgenetic alopecia). |
| Any significant active scalp inflammation, psoriasis, or seborrheic dermatitis that required topical treatment to the scalp, significant trauma to the scalp, or untreated actinic keratosis on the scalp. |
| Any relevant immunological conditions including uncontrolled thyroid disease and systemic/cutaneous lupus. |
| Any males with co-existent androgenetic alopecia (Hamilton Norwood Grade IV or more) and women with female pattern hair loss (Sinclair Score 3 or more). |
| Any other chronic co-morbidity that was not controlled by appropriate medication. |
| Any patients with allergies to either soyabean or peanuts |
| *Prohibited medications* |
| Any topical hair loss treatment in the previous 6 weeks. |
| Any oral janus kinase inhibitors (e.g., tofacitinib, ruxolitinib) in the previous 6 weeks. |
| Any systemic immunosuppressive agent (e.g., cyclosporine, oral corticosteroids, mycophenolate mofetil, azathioprine, methotrexate, or tacrolimus) in the previous 6 weeks. |
| Any topical/oral calcineurin inhibitors or minoxidil medication in the previous 6 weeks. |
| Any intralesional, intra-articular or depot corticosteroid medication in the previous 3 months |
| Any investigational drug treatment (i.e., clinical trial involvement) in the previous 3 months. |
| *General exclusions:* |
| Any history of cutaneous or systemic medical illness or psychiatric disease that would put the patient at risk or interfere with assessments. |
| Any known human immunodeficiency virus or hepatitis B or C positive conditions. |
| Any vaccination with herpes zoster or any live virus in the previous 6 weeks. |
| Hospitalisation or change of chronic concomitant medication in the previous 6 weeks, or any planned hospitalisation during trial involvement. |
| Symptoms of cardiovascular disease or heart related problems. |
| Drug or alcohol abuse. |
| Women who were pregnant or nursing |
| Any condition which, in the opinion of the Investigator, made the patient unsuitable for inclusion. |

### Endpoints

#### Primary

The primary endpoints were derived from Severity of Alopecia Tool (SALT) score ratings, which were evaluated for:

- Clinical responder rate
- Percentage change from baseline
- Dose response
- Percentage change over time
- Overall risk/benefit evaluation

#### Secondary

- Hair growth quantitative measurement (patches)
- Alopecia Areata Symptom Impact Scale (AASIS) scores – *to be reported elsewhere*
- Alopecia Areata Quality of Life Index (AAQLI) scores - *to be reported elsewhere*
- Clinical Global Impression-Improvement (CGI-I) scores
- Clinical Global Impression-Severity (CGI-S) scores
- Immunological markers of efficacy, including: IFNγ, TNFα, IL-4, IL-10, IL-12, IL-13, IL-17, and IL-18 - *to be reported elsewhere*

#### Exploratory

- Investigation of potential factors of interest or sources of variability that impacted the distribution of the magnitude of treatment effect (SALT score)
- SALT score cumulative distribution of treatment response
- SALT score time to response

#### Safety

- AEs
- Physical examination
- Vital signs
- Safety laboratory evaluations
- Prior and concomitant medications

### Compliance

**Table S2.** Compliance classification and weight of cream used during the treatment assessment.

| Assessment of compliance | Placebo<br>(N=32) | STS01 cream |  |  |  |
| --- | --- | --- | --- | --- | --- |
|  |  | 0.25%<br>(N=30) | 0.5%<br>(N=31) | 1%<br>(N=31) | 2%<br>(N=31) |
| **2 months, n (%)** |  |  |  |  |  |
| No answer | 10 (31.3) | 9 (30.0) | 9 (29.0) | 9 (29.0) | 10 (32.3) |
| Non-compliant | 0 (0.0) | 0 (0.0) | 0 (0.0) | 0 (0.0) | 2 (6.5) |
| Minimally | 0 (0.0) | 0 (0.0) | 2 (6.5) | 3 (9.7) | 5 (16.1) |
| Moderately | 2 (6.3) | 8 (26.7) | 13 (41.9) | 9 (29.0) | 6 (19.4) |
| Fully | 20 (62.5) | 13 (43.3) | 7 (22.6) | 10 (32.3) | 8 (25.8) |
| *Weight of cream used<sup>†</sup> [g]* |  |  |  |  |  |
|  | (n=24) | (n=22) | (n=19) | (n=22) | (n=19) |
| Median | 28.75 | 20.60 | 20.70 | 17.31 | 17.20 |
| Min, max | 5.5, 52.0 | 6.2, 50.8 | 8.1, 55.9 | 9.3, 44.1 | 8.9, 36.7 |
| **4 months, n (%)** |  |  |  |  |  |
| No answer | 10 (31.3) | 9 (30.0) | 9 (29.0) | 11 (35.5) | 9 (29.0) |
| Non-compliant | 1 (3.1) | 2 (6.7) | 1 (3.2) | 2 (6.5) | 2 (6.5) |
| Minimally | 0 (0.0) | 0 (0.0) | 4 (12.9) | 3 (9.7) | 6 (19.4) |
| Moderately | 4 (12.5) | 8 (26.7) | 6 (19.4) | 7 (22.6) | 6 (19.4) |
| Fully | 17 (53.1) | 11 (36.7) | 11 (35.5) | 8 (25.8) | 8 (25.8) |
| *Weight of cream used<sup>†</sup> [g]* |  |  |  |  |  |
|  | (n=21) | (n=20) | (n=21) | (n=22) | (n=20) |
| Median | 26.00 | 26.57 | 22.00 | 15.05 | 11.30 |
| Min, max | 3.6, 59.5 | 4.7, 53.4 | 2.4, 54.1 | 0.2, 50.5 | 6.2, 49.7 |
| **6 months, n (%)** |  |  |  |  |  |
| No answer | 9 (28.1) | 9 (30.0) | 6 (19.4) | 8 (25.8) | 7 (22.6) |
| Non-compliant | 2 (6.3) | 1 (3.3) | 1 (3.2) | 4 (12.9) | 7 (22.6) |
| Minimally | 2 (6.3) | 2 (6.7) | 4 (12.9) | 6 (19.4) | 6 (19.4) |
| Moderately | 5 (15.6) | 8 (26.7) | 6 (19.4) | 5 (16.1) | 3 (9.7) |
| Fully | 14 (43.8) | 10 (33.3) | 14 (45.2) | 8 (25.8) | 8 (25.8) |
| *Weight of cream used<sup>†</sup> [g]* |  |  |  |  |  |
|  | (n=23) | (n=24) | (n=26) | (n=25) | (n=23) |
| Median | 32.80 | 20.27 | 21.90 | 14.47 | 12.00 |
| Min, max | 2.1, 60.0 | 0.0, 49.0 | 1.0, 53.3 | 0.0, 52.8 | 1.1, 50.9 |

Non-compliant = up to 25% of days, minimally compliant = up to 50% of days, moderately compliant = up to 75% of days, fully compliant = up to 100% of days
<sup>†</sup> Assumed 70 g weight at start and summarises the weight of cream used between visits.

### Clinical response

**Table S3.** Clinical response, ≥30, 40, 50, 60, and 100% improvement in SALT score after 6 months (LOCF: FAS)

|  | Placebo | STS01 cream |  |  |  |
| --- | --- | --- | --- | --- | --- |
|  |  | 0.25% | 0.5% | 1% | 2% |
| *Improvement* | (N=32)/<br>(n=30) | (N=30)/<br>(n=29) | (N=31)/<br>(n=28) | (N=31)/<br>(n=29) | (N=31)/<br>(n=28) |
| $\geq 30\%$, N (%) | 11 (36.7) | 11 (37.9) | 16 (57.1) | 22 (75.9)** | 17 (60.7) |
| 95% CI | (19.4, 53.9) | (20.3, 55.6) | (38.8, 75.5) | (60.3, 91.4) | (42.6, 78.8) |
| $\geq 40\%$, N (%) | 9 (30.0) | 9 (31.0) | 16 (57.1) | 18 (62.1)* | 15 (53.6) |
| 95% CI | (13.6, 46.4) | (14.2, 47.9) | (38.8, 75.5) | 44.4 (79.7) | (35.1, 72.0) |
| $\geq 50\%$, N (%) | 7 (23.3) | 9 (31.0) | 14 (50.0) | 15 (51.7)* | 14 (50.0) |
| 95% CI | (8.2, 38.5) | (14.2, 47.9) | (31.5, 68.5) | (33.5, 69.9) | (31.5, 68.5) |
| $\geq 60\%$, N (%) | 6 (20.0) | 8 (27.6) | 13 (46.4)* | 15 (51.7)* | 13 (46.4)* |
| 95% CI | (5.7, 34.3) | (11.32, 43.9) | (28.0, 64.9) | (33.5, 69.9) | (28.0, 64.9) |
| **100%**, N (%) | 1 (3.3) | 3 (10.3) | 6 (21.4)* | 5 (17.2) | 7 (25.0)* |
| 95% CI | (0, 9.8) | (0, 21.4) | (6.2, 36.6) | (3.5, 31.0) | (9.0, 41.0) |

CI, confidence interval; FAS, full analysis set; LOCF, last observation carried forward; N, number of patients in the treatment group; n, number of patients with data at baseline and post-baseline (LOCF)
\* $p < 0.05$ ; \*\* $p < 0.01$

### Exploratory analysis

To determine whether any of the baseline demographics or clinical variables could potentially impact the distribution and magnitude of the treatment effect on the SALT score, a regression analysis was performed. None of the variables significantly contributed to the model at the 10% level of significance, and the only statistically significant factor influencing the outcome was the treatment group (Table S4).

**Table S4.**
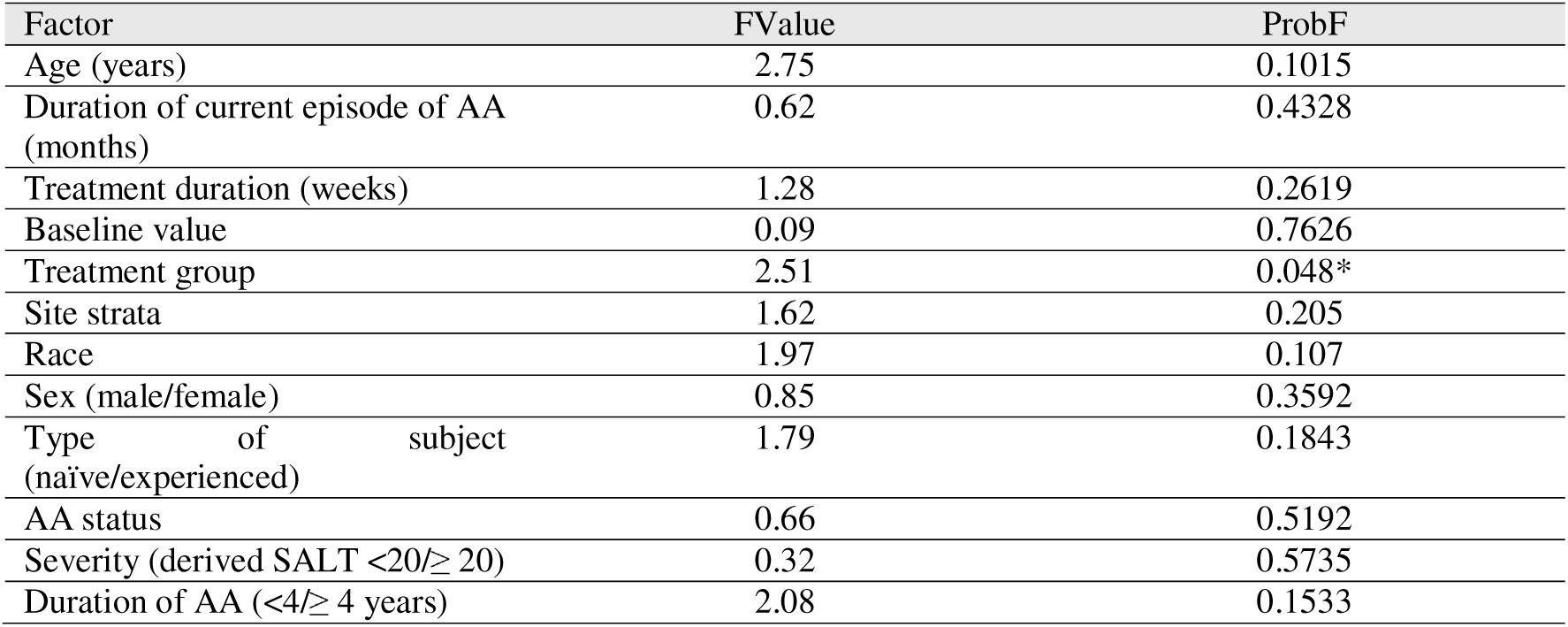
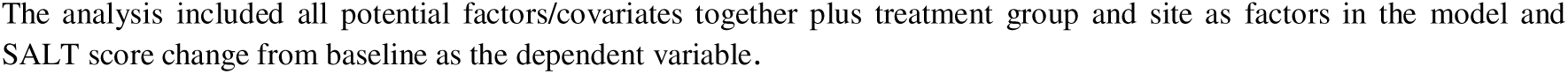
Impact on Outcome: Regression model with all prognostic variables as main effects.

A separate GLM model analysis using AA severity (mild/moderate) at baseline alone as the main effect and treatment*subgroup interaction term was used to quantify the magnitude of the treatment effect across the doses in both subgroups (i.e mild and moderate). Both exhibited similar dose response curves to that of the main analysis albeit with a more gradual improvement gradient with dose particularly for the milder patient group. This differed only quantitatively and was probably a result of the lower starting SALT scores (Figure S2).

**Figure S2.**
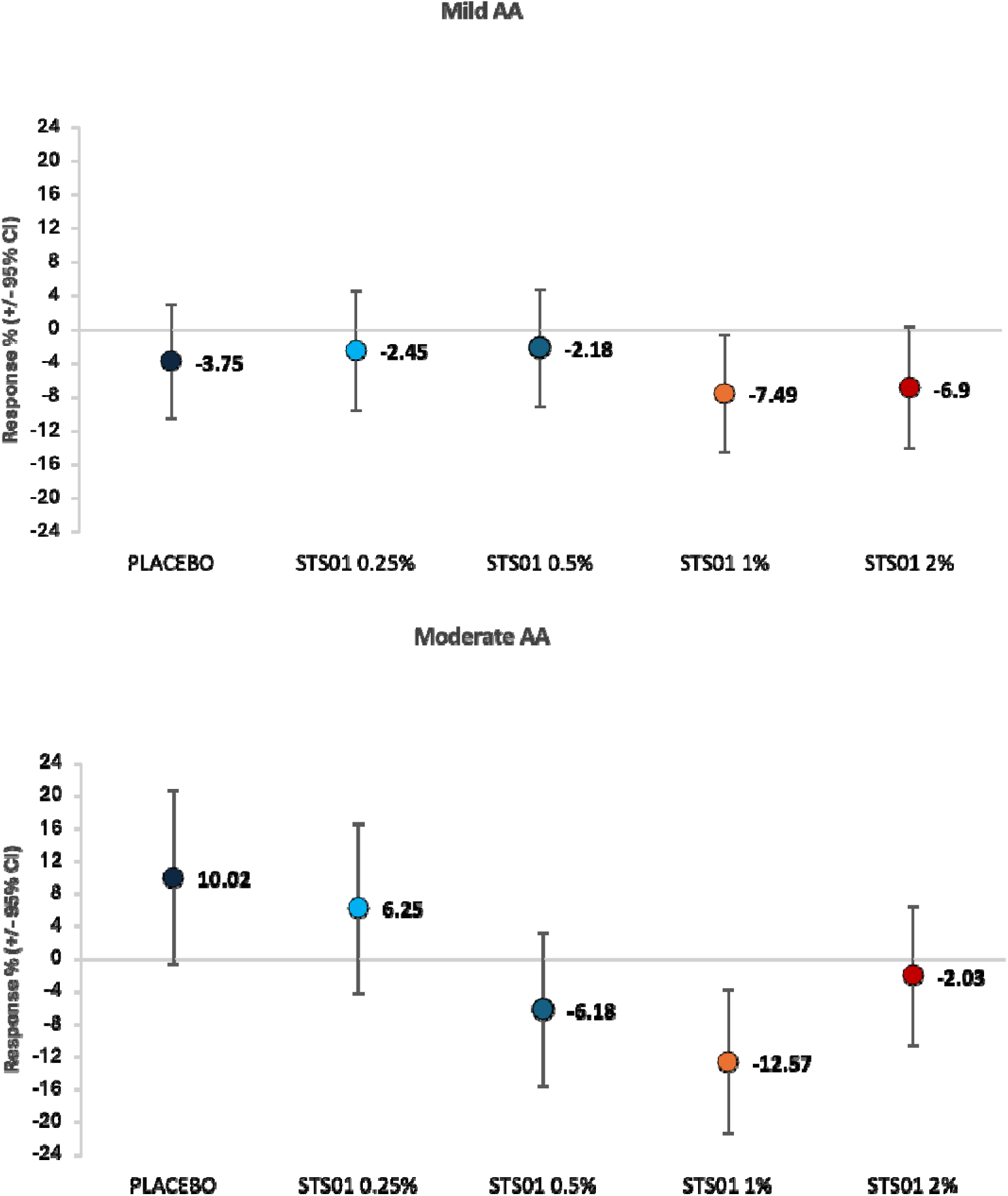
AA Severity: LS mean change from baseline (95% CI) in (A) mild AA and (B) moderate AA. ANCOVA model with Main effects + Treatment*Subgroup interaction

### Risk-benefit analysis

The relationship between efficacy, safety and compliance for each treatment dose group was also investigated at each visit and the end of the treatment period (Visit 5, 6 months) with a 3-spine radar plot.

Each of the 3 spines was displayed by visit with efficacy as mean percentage decrease (maximum improvement=100%) SALT score percentage change from baseline. With safety averaged as the mean incidence of erythema (any redness/irritation reported over as 0=no irritation, 1=mild, 2=moderate, 3=severe) as a proportion of the maximum possible score 3. Compliance was averaged from 0=not answered, 1=non-compliant, 2=minimally, 3=moderately, 4 = fully, as a proportion of the maximum possible score 4. This combination of endpoints could help to support the choice of treatment dose in phase 3 clinical development.

Note that risk-benefit analysis of efficacy and safety are based on crude average scoring.

**Figure S3.**
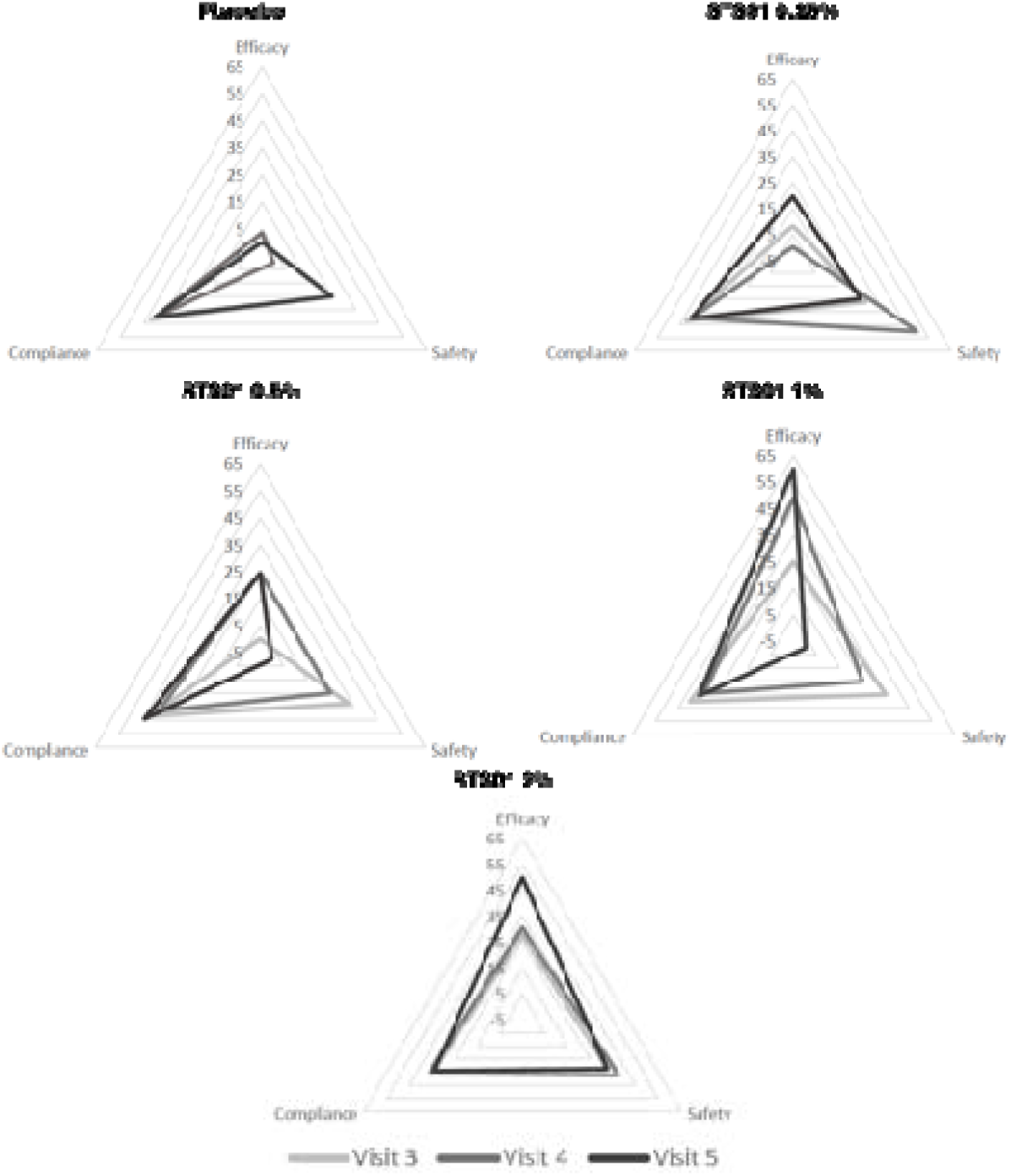
Overall risk-benefit evaluation (FAS) FAS, full analysis set; LOCF, last observation carried forward; SALT, Severity of Alopecia Tool Weighting applied efficacy as mean % decrease (improvement (max=100%)) SALT score change from baseline after 6 months or withdrawal (LOCF); safety as any mean incidence of irritation reported over (weighted for each patient (from worst report over 6 months) as 0=no irritation (0%), 1=mild (33%), 2=moderate (66%), 3=severe (99%); compliance average (weighted for each patient (from worst report over 6 months) 0=not answered (0%), 1=non-compliant (25%), 2=minimally (50%), 3=moderately (75%), 4=fully (100%)

## Notes

### Competing Interest Statement

The authors have declared no competing interest.

### Clinical Trial

NCT06402630/ Eudract 2021-004145-20

### Funding Statement

Study was funded by Soterios LTD

### Author Declarations

The responsible Independent Ethics Committee (London - South East Research Ethics Committee) approved the study protocol (Reference: 21/LO/0851).

